# Ionizing radiation acoustic beam localization: one step towards "proton surgery"

**DOI:** 10.64898/2026.03.07.26347755

**Authors:** Wei Zhang, Ibrahim Oraiqat, Jiyeon Park, Glebys Gonzalez, Yiming Liu, Yaocai Huang, Sarah Dykstra, Lise Wei, Dale Litzenberg, Kyle Cuneo, William Mendenhall, Curtis Bryant, Samuel Jean-Baptiste, Perry Johnson, Issam El Naqa, Xueding Wang

## Abstract

Proton beam therapy (PBT) offers a unique potential for dose conformity to tumors while sparing surrounding healthy tissues. Current PBT accuracy, however, is fundamentally limited by range uncertainties from tissue density variations and anatomical changes, yet no clinically viable methods exist for localizing the dose delivery pulse-by-pulse inside patients during pencil beam scanning (PBS). We developed and clinically demonstrated a first-of-its-kind radiation acoustic beam localization (iRABL) system for real-time tracking PBS trajectory and mapping dose deposition deep in patient’s body during PBT. A clinical-grade compact iRABL system featuring high speed, super-resolution, and high sensitivity was specifically designed for PBT applications. Its clinical feasibility was validated through the first-in-human study on prostate cancer patients, demonstrating the capability for *in vivo* proton dose mapping without interfering with treatment delivery. System performance, including spatial resolution, imaging speed for tracking beam trajectory and temporal dose accumulation, and dosimetric accuracy, was quantitatively characterized using tissue-equivalent phantoms and clinical treatment plans. This iRABL system achieved displacement resolution of 0.1 mm laterally and 0.2 mm axially, exceeding the acoustic diffraction limit by an order of magnitude and surpassing typical proton beam spot sizes. This super-resolution capability, combined with GPU-accelerated image reconstruction and processing, enabled single-pulse detection at a frame rate of 1 kHz, matching the proton system’s pulse repetition rate. Dosimetric validation using clinical M-shaped treatment plans met clinical criteria with gamma index passing rates exceeding 90% at 3 mm/3% tolerance, confirming high accuracy for mapping delivered dose distributions. For the first time, by leveraging the high sensitivity and the high speed of our newly developed iRABL system, we are able to localize proton beam and map the proton dose deposition during PBS with sub-diffraction-limit spatial resolution, pulse-by-pulse imaging speed, and clinical grade accuracy. This capability, which addresses fundamental limitations in current treatment monitoring, holds promise for advancing PBT toward image-guided "proton surgery".

## Introduction

Proton beam therapy (PBT) has become an increasingly important modality in the treatment of cancer, owing to its unique physical characteristics that can enable highly conformal dose delivery [1, 2]. Unlike conventional photon-based radiation therapy, PBT exploits the distinct Bragg peak phenomenon, where the majority of the proton energy is deposited at a specific depth in tissue, beyond which the dose rapidly falls off [3]. This allows for maximal irradiation of the tumor while sparing adjacent healthy tissues and critical structures—a particularly valuable advantage in treating pediatric cancers or tumors near sensitive organs [4]. Achieving the full therapeutic potential of PBT, however, hinges on accurate delivery of the prescribed dose to the precise location of the tumor. Pencil beam scanning (PBS) proton therapy delivers highly conformal dose distributions by scanning narrow, magnetically focused proton spots across the target and is now the preferred and most rapidly expanding delivery technique in proton therapy because it can effectively “paint” complex tumors while sparing nearby organs at risk [5]. Unfortunately, limitations, including anatomical changes, physiological motion, and range uncertainties caused by variations in patient setup or tissue composition, result in the Bragg peak and dose being misplaced—potentially undermining tumor control or increasing normal tissue toxicity [6-9]. One of the key side effects is the loss of distal edge sparing due to these uncertainties, which highly reduce the application of PBT in small targets, such as gliomatosis cerebri [10]. In addition, the robustness of intensity modulated proton therapy plans, which strongly rely on the distal fall-off, will be highly reduced [11].

Despite extensive advances in proton therapy planning and delivery, a fundamental and serious technical gap remains, which is the lack of reliable, real-time, precise imaging method for localizing proton pencil beam and mapping dose deposition inside a patient body during treatment. Especially for proton therapy based on PBS, such a gap prevents cutting-knife, millimetric, and biologically exact dose deposition in the way “surgery” implies [12]. To fill this technical gap and advance PBT toward "proton surgery", we need an imaging technique for localizing proton beam in patient body with high spatial resolution on the level of submillimeter, high detection sensitivity on the level of single proton pulse, and high imaging speed to map the proton dose deposition pulse-by-pulse [13]. Current imaging, such as X-ray CT and MRI, primarily provide anatomical information or post-treatment verification, but do not offer real-time monitoring of actual proton range or dose deposition during irradiation [6]. *In vivo* dosimeters and point detectors, while useful, are often limited to measurements near patient’s body surface or at discrete locations, lack volumetric coverage, and are unable to directly verify proton energy deposition at depth [14]. Prompt gamma imaging [15] and positron emission tomography (PET) [16] can be used with proton delivery, but do not directly report dose.

Our previously demonstrated ionizing radiation acoustic imaging (iRAI) technique, with a unique capability of mapping radiation dose deposition in deep patient body during conventional radiation therapy, represents a potential solution to address this need [17]. Several earlier studies, based on the similar thermo-acoustic or radiation acoustic imaging concept, have also explored the physical principle and basic feasibility for range verification in PBT [18-20]. However, suffering from the limited imaging resolution (on the order of a few millimeters), imaging speed (only show integrated dose), detection sensitivity (large-scale average over tens or hundreds of pulses), and dose mapping accuracy (not quantitative in 3D mapping of the dose), none of the current thermos-acoustic or radiation acoustic imaging systems, including those developed by our lab, can meet the demanding requirements of proton beam localization and dose mapping for advancing PBT toward the goal of “proton surgery” [21-23].

In this study, we developed and clinically demonstrated a first-of-its-kind radiation acoustic beam localization (iRABL) system for real-time tracking PBS trajectory and mapping dose deposition deep in patient’s body during PBT. A clinically ready iRABL system with capability of localizing proton beam delivery *in vivo*, in real time, was specially developed for PBT based on PBS. The clinical feasibility of this system was evaluated on prostate cancer patients. A comprehensive performance validation of this super-resolution iRABL system was conducted on tissue-equivalent phantoms. For the first time, by leveraging the high sensitivity and the high speed of the iRABL system, we were able to localize proton beam and map dose deposition during PBS with unprecedented spatial resolution (0.1 mm, exceeding the acoustic diffraction limit by an order of magnitude and surpassing typical proton beam spot sizes), imaging speed (1 kHz frame rate, enabling single-pulse detection at a frame rate of 1 kHz and matching the proton system’s pulse repetition rate), and accuracy (90% gamma index passing rates at 3mm/3% tolerance, exceeding clinical criteria). Our results highlight the potential of iRABL to address the long-standing and critique limitations of PBT, render a method for treatment guidance leading to high-precision PBT, and pave the way toward “proton surgery”.

## Method

### A volumetric ionizing radiation acoustic beam localization system

To localize the proton dose delivery during the PBT, especially to track the pulse-by-pulse PBS trajectories, a high-speed, high sensitivity and high-resolution volumetric iRABL system was specially designed, as shown in Figure 1A. A 32×32 = 1024 elements 2D matrix array transducer (13961 01002, Imasonics Inc) with a central frequency of 350 kHz and 50% bandwidth was custom-designed to optimize the detection sensitivity to match the spectrum of acoustic signals induced by 7-µs proton pulses. A custom-built 1024-channel preamplifier array (AMP 1024-25-001, Photosound Inc) with 46 dB gain was directly integrated with each element of the 2D matrix array to minimize the potential noise when using cables between the transducer array and the preamplifier array. An additional 80 dB gain, including 24 dB low noise amplifier, 26 dB TGC, and 30 dB preamp output buffer, was set in the acquisition system to fully utilize the dynamic range of the data acquisition system. The output impedance of the preamplifier array was set to match the input impedance of the Verasonics Vantage system (Vantage 256, Verasonics Inc). With a 256-channel data acquisition system, the adjacent four elements were sequentially selected via a 4 to 1 multiplexer. During iRABL imaging, the 2D matrix array transducer is held by a custom built articulated mechanical arm, which provides 6 degrees of freedom. The arm was directly mount to the couch to avoid relative motion during the couch movement for patient alignment. All the control for data acquisition was achieved via a remote PC placed outside of the treatment room connected via wireless internet.

**Figure 1.**
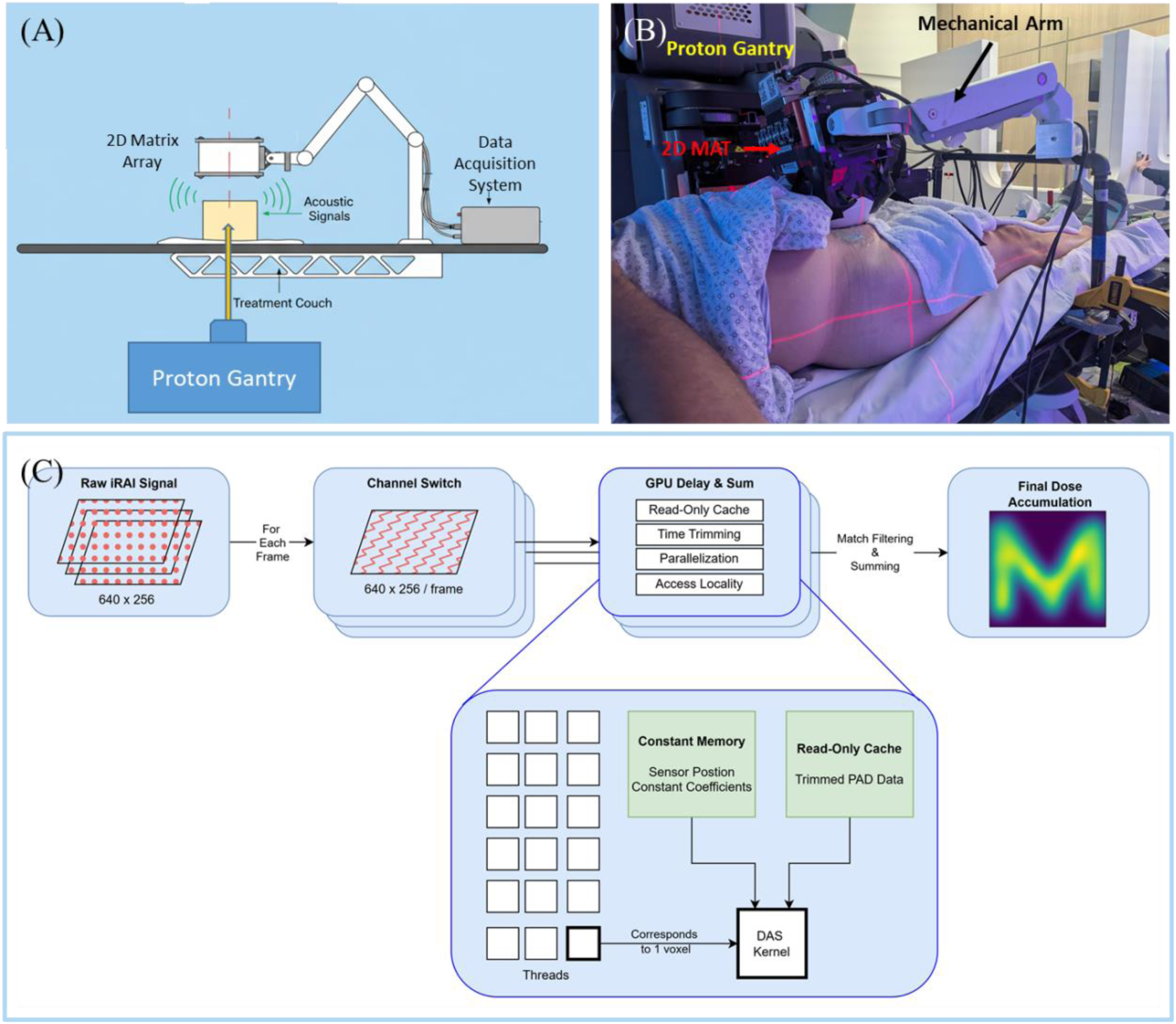
The 3D volumetric ionizing radiation acoustic beam localization (iRABL) system and the GPU acceleration framework. (A) The schematic of the iRABL system working with an IBA Proteus ONE proton beam device. (A) The photograph of the iRABL system setup for patient measurement during PBT. (C) The schematic of the GPU accelerated data processing framework with the delay-and-sum (DAS) image reconstruction algorithm.

### Clinical Trial Design

The clinical feasibility of the proposed system was first evaluated in prostate patients treated with PBT. The clinical trial protocol for this pilot study (UFPTI 2403-PR12: Real-Time Volumetric Ionizing Radiation Acoustic Imaging for In-vivo Proton Treatment Monitoring in Pencil-Beam Scanning, IRB202400573) was approved by the Institutional Review Board (IRB) of the University of Florida. Informed consent was obtained from each patient prior to enrollment. Eligibility criteria included the presence of a tumor in the prostate or prostate plus seminal vesicles without pelvic nodal involvement and a clinical indication to receive PBT. The dose prescription was determined by the treating physician, with fractional doses (relative biological effectiveness of 1.1) ranging from 2.0 to 2.6 Gy over 28-39 fractions. Each patient had three implanted fiducial markers (Gold Anchor, Naslund Medical AB) and a rectal spacer (SpaceOAR, Boston Scientific) for image guidance and anterior rectal wall dose spearing during treatment. PBT was delivered using a compact gantry operated with a synchrocyclotron system (Proteus^®^ ONE, IBA). Patient treatment alignment was verified with daily oblique x-ray image guidance focusing on the fiducial markers, which also served to confirm iRABL system positioning without requiring additional imaging. The iRABL measurement was performed for 3 fractions of a total of 4 patients from October 2024 to September 2025. Prior to positioning the 2D matrix array transducer of the iRABL system, diagnostic ultrasound imaging (M6 with a convex C5-2 probe, MindRay) was performed to identify acoustic windows.

### Study Procedures

Treatment plans were created using treatment planning software (Raystation, RaySearch Laboratories). Before the treatment, the 2D MAT and mechanical arm were mounted to the couch, which was placed on the loading position. At the same time, the iRABL system was initialized and synchronized with the proton system to minimize the total time for the patient in this study. During treatment, the patient was first set up per standard of care procedures and oblique x-ray was used to verify the position of fiducial markers in prostate for position verification. The system setup for performing iRABL measurements on a patient is shown in Figure 1B. To facilitate acoustic coupling, a water-filled balloon, with its surface coated with ultrasound coupling gel, was directly attached to the surface of the array. The other side of the balloon was placed to make light contact with the patient’s abdominal skin, limiting potential deformation of patient’s body. The 2D MAT was then placed on the top of the patient’s prostate with the acoustic path pre-verified via diagnostic ultrasound imaging. The relative position between 2D MAT and patient body was recorded by a 3D structure camera (L215u, LIPS Corporation). The total distance between the matrix array and the skin was set to 5 cm. Before delivering the first portion of the dose, the patient’s position was reconfirmed using pre-established markers and second oblique x-ray to mitigate any potential changes related to the placement of the matrix array. The proton beam from the left side of the patient was then delivered to the target. After the first beam angle was delivered, the couch rotated 180 degrees to allow the proton beam delivery from the right side of the patient body. The third oblique X-ray was taken to make sure that rotation of the couch did not change the target position. Then, the second beam angle, which contains half of the dose, was delivered to the target. Once the second portion of the proton beam was delivered, the couch was moved back to the loading position to remove the 2D MAT from the patient’s abdomen. All the proton beam pulse induced acoustic signals were acquired pulse-by-pulse during the treatment. The acquired signals were processed to localize the dose distribution. With the position of the 2D MAT relative to the patient geometry recorded by the 3D structure camera, imaging fusion between simulation CT and iRABL dose distribution was performed. The alignment between treatment plan and iRABL measured dose distribution was evaluated with 60% and 80% iso dose lines.

### GPU acceleration

By leveraging GPU acceleration, our iRABL system works in a pulse-by-pulse mode with an imaging frame rate of 1 kHz (i.e., the proton pulse repetition rate) and meaning that both data acquisition and 3D image reconstruction after each proton pulse can be completed within 1 ms of proton pulse interval. The schematic of the GPU acceleration is shown in Figure 1C. The kernel configuration is optimized with a block size of 256, which maximizes GPU occupancy and balances register usage. The detected raw acoustic signals are first processed into a tensor of shape 640 × 32 × 32 for each frame. Each frame is then reconstructed individually using a GPU-accelerated delay-and-sum (DAS) kernel into a 3D volume covering the region of interest. For efficient computation, the raw acoustic signal data are flattened and stored in the GPU’s read-only cache, allowing all threads to access them with spatial and temporal locality considered. The sensor positions and other constant parameters, including the sampling frequency and half-aperture threshold, are stored in constant memory, as these coefficients remain identical for all voxels. Each GPU thread computes the reconstruction for a single voxel, writing results into a shared GPU array that is later transferred back to the CPU for aggregation. During each kernel run, the time range is trimmed based on the time of flight to the region of interest, minimizing unnecessary memory access for each voxel. Image reconstructions were performed on two GPU cards, respectively. One of them is an NVIDIA A100 with 80 GB RAM, while the other is an NVIDIA 3090 GPU with 24 GB of GDDR7 RAM that is much lower in cost. The outcomes from them were compared. The evaluation of the image reconstruction speed also considered different sizes of the region of interest.

The speed of GPU-accelerated image reconstruction was evaluated across 100 pulses, with results for different settings shown in Table 1. Powered by an NVIDIA A100, iRABL can process a 5 cm × 5 cm × 5 cm region of interest with a 0.5 mm grid size in 0.5 ms, sufficient for single proton pulse position tracking. Even working with the low-cost NVIDIA RTX 3090 GPU, single proton pulse positions can also be tracked in less than 1 ms for the same region and grid size. Using the current settings, neither the A100 nor RTX 3090 alone can process a larger 10 cm × 10 cm × 10 cm region (typical of the largest planning target volumes in PBT) within one pulse interval of 1 ms, which, however, can be achieved by using a GPU array or a more powerful GPU.

**Table 1.** Gamma index analyses for iRABL tracked dose distribution for PBT.

|  | NVIDA A100 80 GB | NVIDA RTX 3090 24GB |
| --- | --- | --- |
| $5 \text{ cm} \times 5 \text{ cm} \times 5 \text{ cm}$ with 0.5 mm grid size | Avg = 0.42 ms; STD = 0.01 ms; Median = 0.42 ms | Avg = 0.49 ms; STD = 0.40 ms; Median = 0.42 ms |
| $10 \text{ cm} \times 10 \text{ cm} \times 10 \text{ cm}$ with 0.5 grid size | Avg = 4.01 ms; STD = 0.01 ms; Median = 4.01 ms | Avg = 4.09 ms; STD = 1.06 ms; Median = 3.90 ms |

### Super-resolution beam localization algorithm

A beam localization algorithm was developed for the iRABL system, by utilizing the pre-calibrated beam profile from the commissioning database. Working in the pulse-by-pulse imaging mode, each iRABL imaging frame only contains the dose from a single proton pulse, and a match filtering method can be applied to localize the range of each proton pulse in a single iRABL frame. The goal of match filtering is to localize the proton beam by finding the location (x0, y0, z0) where the 3D proton dose profile best matches with the iRABL image. If the R(x,y,z) is the iRABL image, and h(x,y,z) is the known beam profile of a certain energy proton beam, the match filter output map M(x,y,z) is computed by the following convolution:

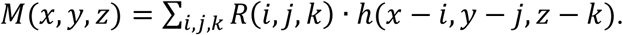

This operation produces a 3D map reflecting the similarity between the beam profile and the iRABL image as a function of the location in 3D space. After matched filtering, the location leading to the maximal similarity in the output map M is used to determine the true range of the proton beam. Once the true range of the proton beam is determined for each iRABL frame, a known proton beam profile will be used to replace the delay-and-sum reconstructed image of each frame, which is defined as a super-frame. The total accumulated proton dose distribution from a treatment plan will then be formed with the accumulated super-frames from all the proton pulses. Following a similar idea of super-resolution ultrasound localization microcopy [24-26], this super-resolution iRABL method can localize the proton beams with a spatial resolution beyond the diffraction limit. The detailed workflow of iRABL for real-time data processing during proton beam delivery is shown in Figure 2.

**Figure 2.**
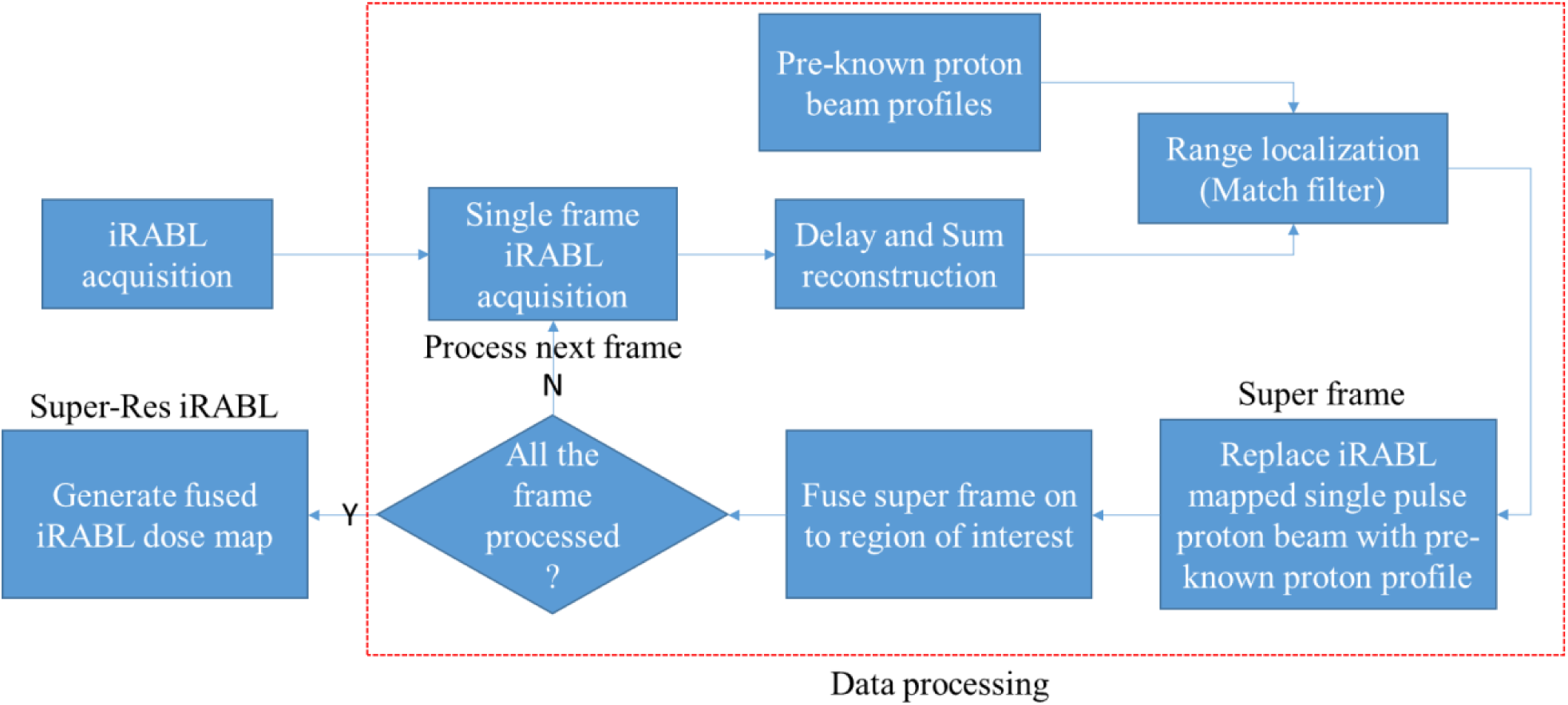
The workflow of iRABL for real-time data processing during proton beam therapy.

### Displacement resolution calibration

To evaluate the performance of the iRABL system in detecting the smallest proton PBS step, a homogenous phantom (15 cm×15 cm×17 cm) made with solidified oil was used. A CT scan of the phantom was acquired on a dedicated simulator (Brilliance BigBore scanner, Philips) with verified Hounsfield Unit to relative stopping power calibration for proton treatment planning. Two treatment plans with different beam scanning trajectories were created to calibrate the displacement resolution in lateral and axial directions, respectively. The treatment plan for lateral displacement resolution calibration consisted of 13 spots, each delivering 12 MU at a beam energy of 120 MeV. The physical coordinates of each proton spot at the isocenter plane are shown in Table 2, with a total lateral span of 10.4 mm and the minimum displacement of 0.1 mm between adjacent spots. The treatment plan for axial displacement resolution calibration consisted of 10 spots positioned at the center of the XY plane (corresponding to the patient’s left and right axis in supine position), each delivering 12 MU. The axial position of each spot (corresponding to the patient’s anterior- posterior axis in supine position) was precisely controlled via beam energy ranging from 120 MeV to 124 MeV with variable energy steps and a smallest step of 0.1 MeV. The energy and axial coordinate of each spot are shown in Table 3. The experimental setup for displacement resolution calibration is shown in Figure 3A. The 2D MAT was held by a mechanical arm to have the detection surface facing downward attached to the top surface of the solidified oil phantom and coupled with ultrasound gel. The center of the phantom was aligned with the iso center of the treatment room. The proton beam was delivered from with a gantry angle of 180 degrees in the posterior-to-anterior direction. Beam alignment with the axial direction of the 2D MAT was verified using oblique x-ray images. Three independent measurements of each treatment plan were performed for statistical analysis.

**Figure 3.**
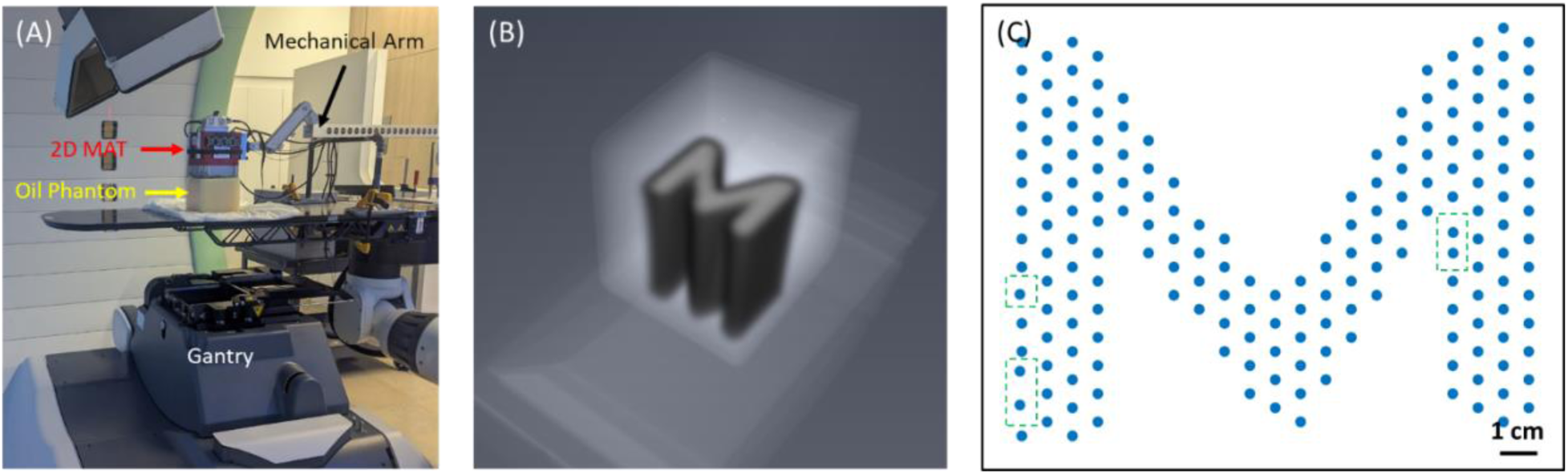
Experimental setup and M-shaped dose distribution treatment plan. (A) Experimental setup for resolution evaluation and beam scanning trajectory tracking. (B) 3D dose distribution for M-shaped treatment plan. (C) The PBS position for M-shaped dose distribution treatment plan of PBT. The green dashed boxes indicate non-uniform distributed spots.

**Table 2.** The physical coordinates of 13 spots for lateral displacement resolution calibration.

|  |  |  |  |  |  |  |  |  |  |  |  |  |  |
| --- | --- | --- | --- | --- | --- | --- | --- | --- | --- | --- | --- | --- | --- |
| X-coordinate [mm] | -6.6 | -3.6 | -1.6 | -1.5 | -0.3 | -0.1 | 0 | 0.1 | 0.3 | 0.8 | 1.8 | 2.8 | 3.8 |
| Y-coordinate [mm] | 0 | 0 | 0 | 0 | 0 | 0 | 0 | 0 | 0 | 0 | 0 | 0 | 0 |
| Spot MU | 12 | 12 | 12 | 12 | 12 | 12 | 12 | 12 | 12 | 12 | 12 | 12 | 12 |

**Table 3.** The physical coordinates and beam energies of 11 spots for axial displacement resolution calibration.

|  |  |  |  |  |  |  |  |  |  |  |
| --- | --- | --- | --- | --- | --- | --- | --- | --- | --- | --- |
| X-coordinate [mm] | 0 | 0 | 0 | 0 | 0 | 0 | 0 | 0 | 0 | 0 |
| Y-coordinate [mm] | 0 | 0 | 0 | 0 | 0 | 0 | 0 | 0 | 0 | 0 |
| Beam Energy [MeV] | 120 | 120.1 | 120.3 | 120.6 | 121 | 121.5 | 122 | 122.5 | 123 | 124 |
| Spot MU | 12 | 12 | 12 | 12 | 12 | 12 | 12 | 12 | 12 | 12 |

### Dynamic proton spot trajectory tracking and validation

As each proton PBS spot is delivered, the iRABL system captures the induced photoacoustic signals and reconstructs the spatial location in vivo, enabling real-time tracking of PBS trajectories and dose delivery. To verify this capability, a treatment plan for a 3D M-shaped target was designed to fit within the solidified phantom, as shown in Figure 3A. The target dimensions were 10 cm in width (patient left-right), length (superior-inferior), and depth (anterior-posterior). The plan was robustly optimized to deliver dose to the 3D M-shaped target using clinical treatment planning parameters, including 3.5% density uncertainty and standard settings for energy layer spacing and spot spacing (scale parameters of 1). Following robust optimization, the initial plan consisted of 30 energy layers ranging from 75 to 150 MeV. To enable detailed analysis of spot localization, spatial resolution, and dose accuracy, three representative energies were selected: low (110.2 MeV), medium (123.5 MeV), and high (141.1 MeV). The monitor units (MU) per spot were set to 12 MU for all selected energy layers. For each energy group, two treatment plans with pencil beam scanning trajectories oriented anterior-posterior and left-right directions were created. The treatment plans were created following the same method previously used for displacement resolution calibration. The treatment plan was optimized to simulate a clinical treatment scenario by targeting a volumetric M-shaped region, accounting for the spot size and spot spacing characteristics of each energy. Depending on the beam energy and corresponding spot size, 130, 150, and 225 spots were delivered, with 12 MU using the Proteus^®^ ONE system. The experimental setup was identical to that used for spatial resolution calibration. Two different experiments were performed using the M-shaped treatment plans to evaluate both the trajectory tracking of the PBS and the temporal dose accumulation monitoring. A representative result of M-shaped dose distribution and corresponding PBS trajectories is shown in Figure 3B and 3C. For statistical analysis, five independent measurements were performed for each treatment plan.

To assess the tracking of PBS trajectory, the radiation-induced acoustic signals were continuously acquired during the proton beam delivery and then processed pulse-by-pulse as described by the workflow in Figure 2. Localization algorithm was applied to each proton beam pulse to extract the proton beam location. The pulse-by-pulse proton beam locations tracked by iRABL formed an imaged trajectory of the pencil beam which was then compared with the beam trajectory in the treatment plan. In the meantime, the pre-calibrated beam profile for corresponding beam energy was used to replace the delay-and-sum reconstructed image of each frame to generate a super-frame iRABL image. The temporal fused super-frame image presents the temporal dose accumulation of the PBS processing during the proton beam delivery. After all the pulses were delivered, the accumulated dose was then compared with treatment plan to evaluate the accuracy of the super-resolution iRABL method for localizing the dose distribution during PBT. In the meantime, the results were also compared with those from conventional iRAI method without using the super-resolution localization algorithm.

## Result

### First in human study of iRABL for PBT

As shown in Figure 4A, 4D and 4G, the predicted proton dose distribution from two lateral beams was overlaid onto the CT scan and displayed in an axial, sagittal and coronal planes, respectively. The dose was normalized to the fractional prescribed dose of 2.5 Gy (RBE = 1.1). The iRABL measured dose from the combined two angles acquired during the treatment is shown in Figure 4B, 4E and 4H. iRABL successfully visualized high-dose region where Bragg peaks were deposited within the planning target volume. However, the low-dose regions proximal to the Bragg peak deposition were not mapped due to the limited field of view of the 2D MAT. Comparing the iRABL-measured and predicted dose distributions, the isodose lines show good overall agreement (Figure 4C, 4F and 4I). To quantitatively assess the accuracy of iRABL dose mapping, the 60% and 80% isodose lines were evaluated. Both the 60% and 80% isodose lines from the iRABL measurement matched well with the treatment plan. Additionally, the posterior aspect of the prostate at the prostate-rectal spacer interface showed reduced dose mapping accuracy compared to the target region. This is likely due to the pubic bone blocking the acoustic signal propagation from the prostate to the 2D MAT.

**Figure 4.**
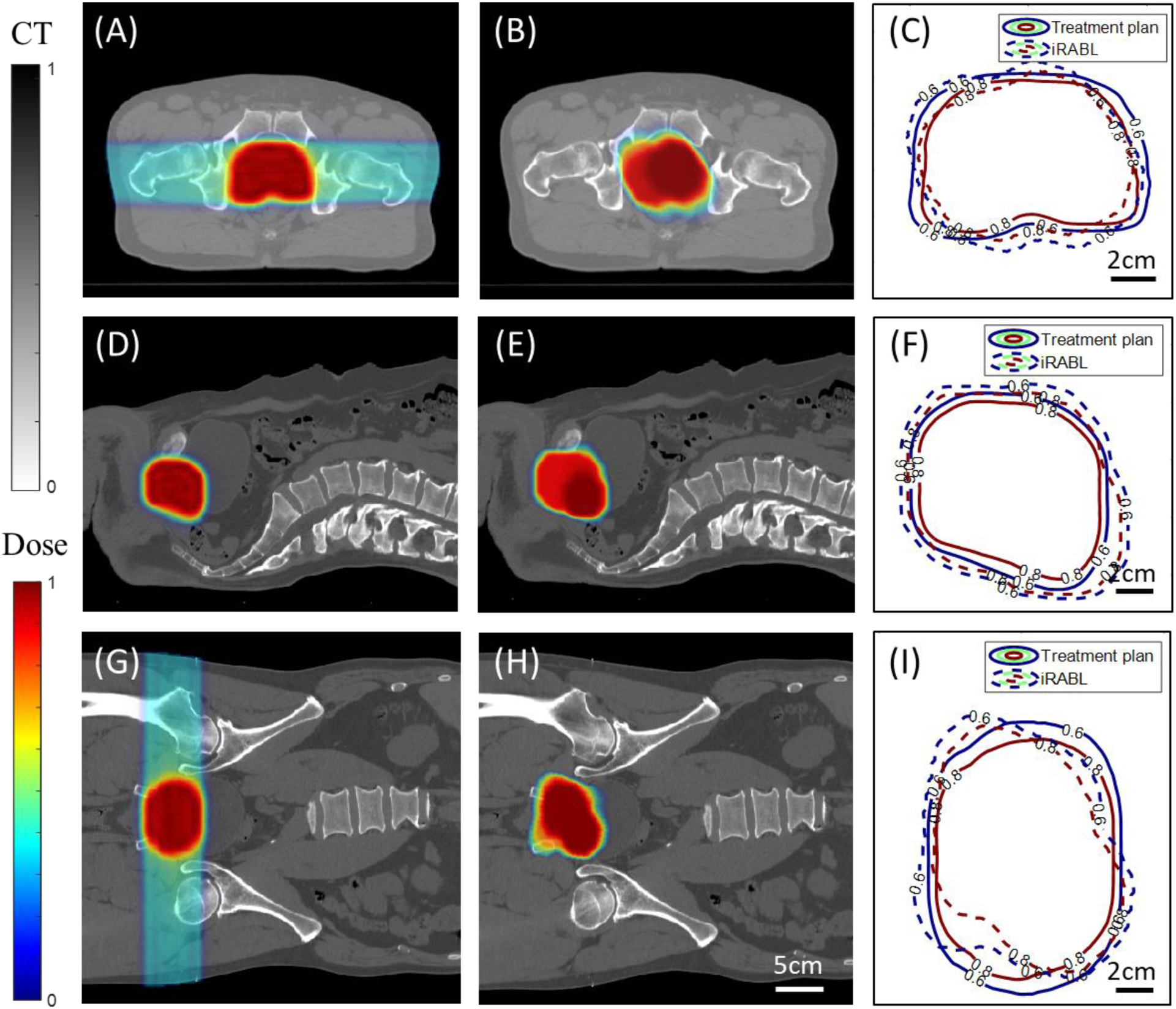
Volumetric dose mapping for proton beam delivery deep into patient prostate during cancer treatment. (A) The dose distribution of the axial plane of the proton beam treatment plan for a prostate cancer patient fused onto the CT scan anatomy structure. (B) The iRABL image of the proton beam dose delivery fused onto the CT anatomy structure with the same position as (A). (C) The 60%, and 80% isodose lines of the axial plane in the iRABL image and the treatment plan. (D)The dose distribution of the sagittal plane of the proton beam treatment plan for a prostate cancer patient fused onto the CT scan anatomy structure. (E) The iRABL image of the proton beam dose delivery fused onto the CT anatomy structure with the same position as (D). (F)The 60%, and 80% isodose lines of the sagittal plane in the iRABL image and the treatment plan. (G) The dose distribution of the coronal plane of the proton beam treatment plan for a prostate cancer patient fused onto the CT scan anatomy structure. (H) The iRABL image of the proton beam dose delivery fused onto the CT anatomy structure with the same position as (G). (I)The 60%, and 80% isodose lines of the coronal plane in the iRABL image and the treatment plan.

### Displacement resolution

The lateral and axial displacement resolution of the IRABL system were quantified by measuring the distance between the Bragg peak locations with different beam delivery positions. By utilizing the super-resolution beam localization algorithm, the Bragg peak location for each pulse was extracted from each iRABL frame reconstructed from DAS algorithm. As shown in Figure 5A, iRABL measurement can resolve the lateral displacement of the proton beam lateral scanning with high accuracy. The mean error and the maximum error of localizing the beam position along the lateral direction are 0.04 mm and 0.25 mm, respectively, with a standard deviation (STD) of 0.14 mm. The lateral scanning of the proton spot with the smallest step size of 0.1 mm can be distinguished successfully by iRABL measurement. Figure 5B shows the iRABL measured axial displacement in relevant to the proton beam depth position change calculated in a treatment planning system. The iRABL measurement can distinguish the displacement of proton beam Bragg peak position along with the beam energy change with high accuracy. The mean error and the maximum error of localizing the beam displacement along the axial direction are 0.07 mm and 0.25 mm, with a STD of 0.05 mm. The minimal axial displacement detectable by iRABL is 0.2 mm, which corresponds to the proton beam travel range difference between 120 MeV and 120.1 MeV in the phantom. This 0.1 MeV energy step represents the typical energy increment tested, as described in the method section. A relatively big variation between the iRABL measured displacement and the ground truth calculated from the treatment planning system is noticed at the beam energy level of 121.5 MeV, which is mostly likely due to the air bubble in the acoustic path between the proton beam deposited position and the 2D MAT. The displacement resolution quantified for iRABL along both the lateral direction (0.1 mm) and axial direction (0.2 mm) are an order of magnitude beyond the diffraction limit of about 2 mm (half the acoustic wavelength at the 350 KHz center frequency of the transducer). It is also worth noting that the displacement resolution quantified here are limited by the scanning step size of the proton machine, not by the imaging system powered by super-resolution beam localization algorithm.

**Figure 5.**
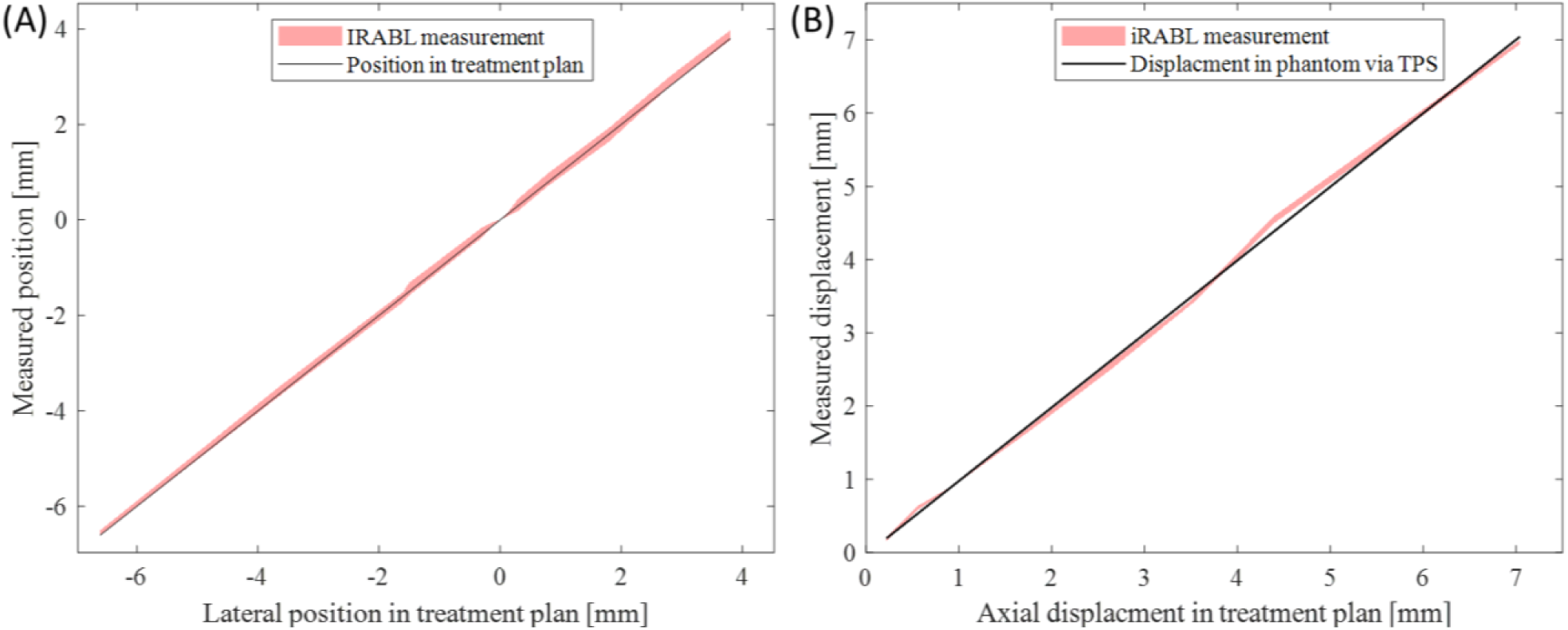
The iRABL measured lateral position and axial displacement of the PBS versus the scanning step size set in the treatment plan. (A) The iRABL measured lateral position of the delivered proton beam versus the settings in treatment plan. (B) The iRABL measured axial displacement of the delivered proton beam versus the settings in treatment plan. The pink line shows the iRABL measurements with the STD presented by shadow, and the black line shows the treatment plan.

### Dynamic proton spot trajectory tracking and validation

With the GPU powered high speed beam localization capability, a frame rate of 1000 Hz, which is the same as the pulse repetition rate of the proton machine used, was achieved for tracking the proton beam trajectory during PBT. A representative result from iRABL showing high-speed pulse-by-pulse proton beam trajectory tracking in a soft tissue phantom is in Supplementary Video 1, where a vertically scanned M-shape treatment plan was successfully imaged. Figure 6 shows the final imaging result from this proton beam tracking experiment presented in Supplementary Video 1. As shown in Figure 6A, the vertically scanned M-shape treatment plan trajectory was tracked by the iRABL system pulse-by-pulse. The M-shape dose distribution formed by 21 individual scanning columns can be seen clearly in the iRABL image. In Figure 6B, the iRABL tracked PBS trajectory are overlayed onto the treatment plan showing the planned scanning spots. For each scanning spot, the iRABL tracked beam locations (white points) are associated with multiple individual proton pulses which are sparsely distributed around the planned scanning spot center marked by the red dot. In the 1^st^ column from the left and the 4^th^ column from the right in Figure 6B, we can see a few spots marked with green dashed box, which were non-uniform distributed in treatment plan, the positions of these spot can still clearly distinguished by iRABL with high accuracy. The observed discrepancy between iRABL measurements and the treatment plan stems from phantom-related uncertainties rather than iRABL system performance. Specifically, minor variations in the solidified oil phantom’s thickness and surface flatness during the experiment, coupled with CT slice thickness limitations that prevented precise coplanar visualization of all planned spot positions, resulted in some planned spots not being presented at the intended plane. This introduced spatial misalignments in the comparison between the planned and measured spot distributions.

**Figure 6.**
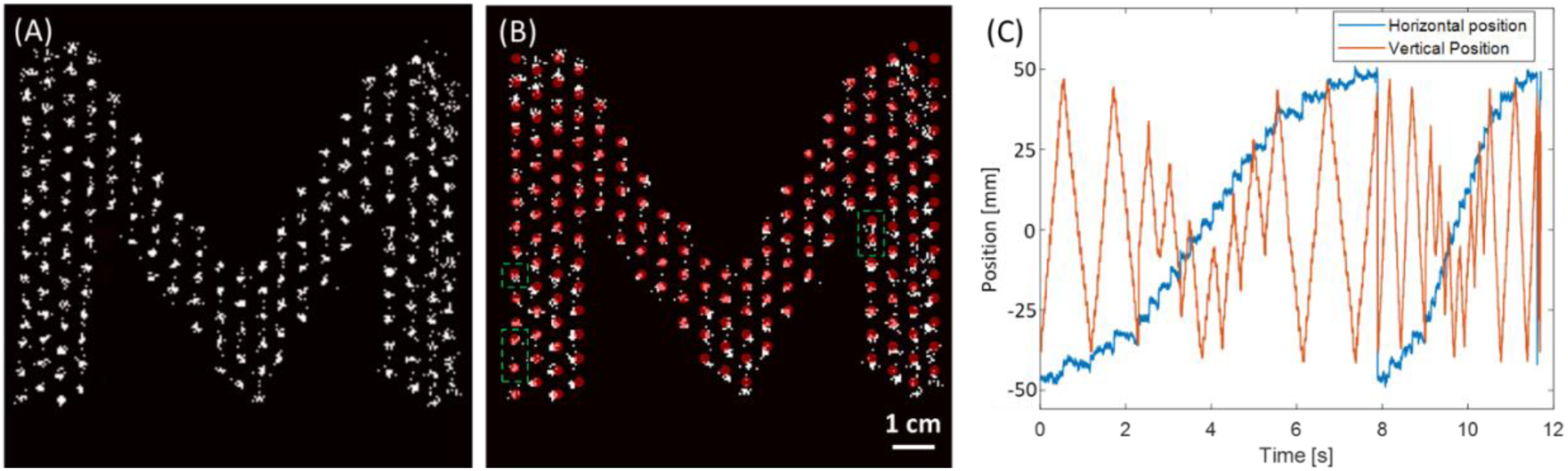
iRABL tracked proton beam scanning trajectory of a vertically scanned M-shape treatment plan. (A) The iRABL tracked PBS trajectory. (B) The iRABL tracked PBS trajectory (white points) overlayed onto the treatment plan (red spots). The green dashed boxes indicate non-uniform distributed spots. (C) The iRABL tracked the pencil beam position as shown by the horizontal and vertical positions over time.

Figure 6C shows the iRABL-tracked proton spot positions over time along the scanning pattern. The vertical axis represents spot scanning along the patient’s left-right direction, corresponding to the fast-scanning axis shown in Figure 6A. The horizontal axis represents spot scanning along the patient’s superior-inferior direction, corresponding to the slow-scanning axis shown in Figure 6A. As seen in Figure 6C, the proton beam scanned in a zigzag pattern along the M-shaped trajectory three times to deliver the planned dose, alternating between the fast and slow scanning axes. The delivered pulses (proportional to dose) for these three scans accounted for approximately 70%, 25%, and 5% of the total delivered dose, respectively. These findings from iRABL imaging align with the delivery characteristics of the Proteus® ONE system (IBA), which typically delivers three bursts per energy layer to ensure monitor unit (MU) accuracy and prevent overdose in synchrocyclotron-based delivery [27]. For delivery efficiency and safety, proton pulses are not delivered to every spot during each scan. Spots requiring relatively low MU can be skipped or delivered in subsequent bursts as part of the spot-based MU modulation strategy employed in synchrocyclotron systems. Once the robustly optimized treatment plan is established with predetermined spot positions and MU values, the delivery system adjusts pulse sequences and burst patterns in real time through feedback control based on cyclotron conditions. This can result in variable burst distributions even when delivering the same treatment plan across multiple fractions. Facilitated by the iRABL tracking system, we successfully imaged the actual trajectory of delivered proton beams during treatment in real time with unprecedented performance: high temporal resolution (single 1-ms pulse interval), high sensitivity (single-pulse detection), and high spatial resolution (0.1 mm lateral and 0.2 mm axial). This capability has not been achieved by any other dosimetry method.

### Temporal dose accumulation

The temporal dose accumulation in the phantom over the delivery time of around 12 seconds was continuously mapped by iRABL. With pulse-by-pulse temporal resolution and sensitivity, iRAI was able to map the temporal dose accumulation with a frame rate of 1 kHz, as shown in Supplementary Video 2. The montage slides in Figure 7A show the time-dependent dose mapping results with a 2-second interval, which demonstrates a gradually formed M-shaped dose distribution over the 12-second treatment. The result for showing the temporal beam and accumulated dose of PBS is shown in Supplementary Video 2. The maximum intensity projection of 3D dose distribution at 3 different planes (coronal, axial and sagittal) over time with PBS during PBT is shown in Supplementary Video 3.

**Figure 7.**
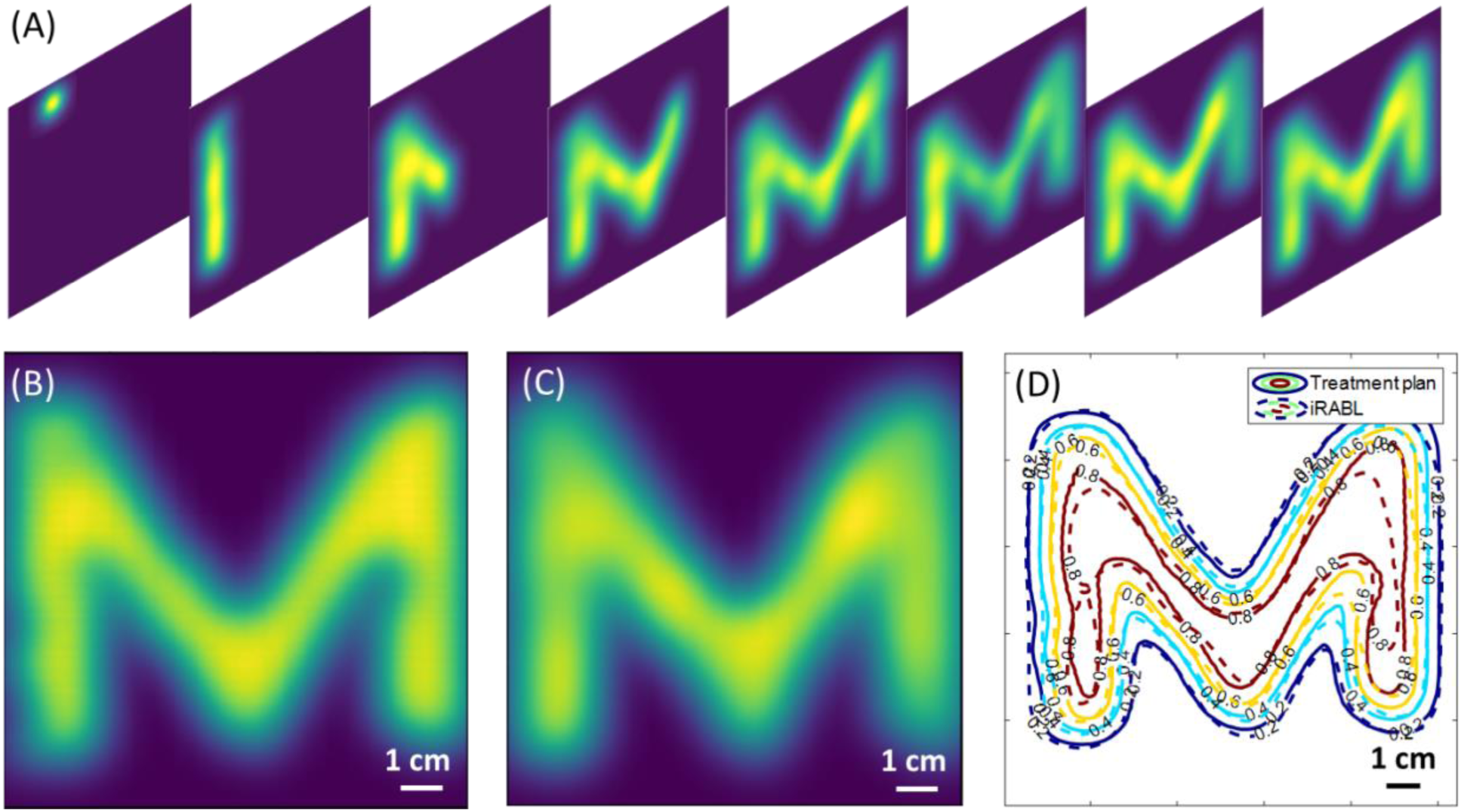
The iRABL imaged temporal dose accumulation. (A) The temporal dose accumulation at different time points imaged by iRABL during the proton beam delivery of an M-shape treatment plan. (B) The cross section of the M-shape treatment plan. (C) The iRABL mapped dose delivery at the same cross section of (B). (D) The isodose lines of the M-shape treatment plan vs the iRABL imaged dose distribution.

### Quantitative evaluation

To further quantify the accuracy of iRABL-tracked the dose distribution during PBT, isodose line analysis [28] and gamma index evaluation [29], both clinical standard methods, were used to compare the iRABL tracked dose distributions with the original treatment plan. As shown in Figures 7B and C, high agreement was observed between the treatment plan as the ground truth and the iRABL-mapped dose distribution at the end of the 12-second treatment delivery. Figure 7D shows the 20%, 40%, 60% and 80% isodose line comparisons between the treatment plan and the iRABL measurements, further demonstrating the accuracy of iRABL in proton dose mapping. Global gamma index with a minimum isodose threshold of 10% was performed to quantify the combination of distance to agreement (DTA) and relative dose difference (DD) between the iRABL image and the treatment plan in 3D. Given the system’s displacement resolution is less than 0.5 mm, passing criteria was set at clinical standard of 3 mm/3% DTA/DD for the iRABL measurement versus the treatment plan. As shown in Table 4, based on independent measurements on 6 different treatment plans, iRABL, powered by the super-resolution beam localization algorithm, achieved an average gamma passing rate higher than 90% at 3mm/3% criteria. This result met clinical standards despite the M-shaped dose distribution featuring steep dose gradients from the periphery toward the center. For comparison, the same treatment plans were also analyzed by the conventional iRAI method with conventional DAS image reconstruction without involving super-resolution beam localization concept [22, 30, 31]. Using the same passing criteria of 3 mm/3% DTA/DD, conventional iRAI achieved gamma index in the range of 55% to 60% for these 6 treatment plans, with an average of 57%. This comparison further demonstrates the substantial improvement in accuracy in localizing the proton beam delivery and mapping the delivered dose achieved by the advanced iRABL method.

**Table 4.** The data processing performance of GPU accelerated iRABL.

| 3mm/3% | 141.1 MeV |  | 123.5 MeV |  | 110.2 MeV |  |
| --- | --- | --- | --- | --- | --- | --- |
|  | Vertical | Horizontal | Vertical | Horizontal | Vertical | Horizontal |
| Conventional | 58.53% | 54.91% | 57.13% | 59.91 | 55.21% | 57.66% |
| iRABL | 73.42% | 95.02% | 90.13% | 96.78% | 87.75% | 91.81% |

## Discussion

While our previously demonstrated iRAI technique offers a unique capability to map radiation dose deposition deep within the patient’s body during conventional radiation therapy, several limitations restrict its applicability for precision proton beam localization [17, 30]. Earlier studies exploring the potential of thermos-acoustic imaging or radiation acoustic imaging for range verification in proton beam therapy, despite showing proofs of principle and basic feasibility [18-20], cannot address the demanding requirements of real-time, pulse-by-pulse, and “pinpoint” beam localization and dose mapping for achieving image-guided “proton surgery”. The goal of this study is to develop and validate a clinically implementable technique to address the long-standing critical need for reliable, real-time, and high-accuracy methods to track pencil beam scanning trajectory and map the dose deposition inside patient’s body during PBT. To achieve this, we demonstrated a clinical-grade compact iRABL system featuring a custom-designed 2D MAT with high sensitivity, high speed, and super-resolution capability, specifically tailored to the unique requirements of PBT. The 2D MAT was optimized with a central frequency matched to the frequency spectrum of the acoustic signals generated by 7-µs proton pulses. The element size of the 2D MAT was selected to balance detection sensitivity with practical operation. A fully integrated and specially designed low-noise multi-channel preamplifier board largely enhanced the system’s sensitivity for detecting proton-induced acoustic signals at the single-pulse level, achieving sufficient signal-to-noise ratio without need of signal averaging. Powered by GPU-accelerated parallel computing and an optimized image reconstruction algorithm, the system could process the acquired acoustic signals from each proton pulse within the pulse interval of 1 millisecond, facilitating real-time pulse-by-pulse beam localization with submillimeter accuracy in both lateral and axial directions during PBT delivery.

The first-in-human study on prostate cancer patients demonstrated that iRABL can image proton beam delivery in real time during clinical PBT without interfering with treatment delivery. The overall dose distribution tracked by iRABL showed high agreement with the treatment plan, indicating that iRABL is a feasible and practical technique for imaging proton beam delivery in clinical settings. Prostate PBT presents a particularly challenging scenario for *in vivo* dosimetry, as relatively high-energy proton beams delivered from lateral beam angles traverse thick femoral bones to reach the deep-seated tumor, creating relatively high range uncertainty. Acoustic signal detection could be impeded in some regions by the pubic bone and air pockets in the bowel. Despite these challenges, our iRABL system successfully mapped the dose deposition within the prostate target with optimization of the 2D MAT angle and placement. For treatment sites at shallower depths or in predominantly soft tissue regions—which represent less challenging acoustic environments—iRABL is expected to provide even more precise mapping of delivered dose distributions across a broader range of clinical PBT cases.

The performance of this iRABL system was comprehensively and quantitatively validated using soft-tissue phantoms and clinical treatment plans. By tracking proton beam delivery with precisely controlled lateral and axial displacements, the super-resolution iRABL achieved displacement resolutions (i.e., minimum detectable motion steps) of 0.1 mm laterally and 0.2 mm axially. This unprecedented spatial resolution for proton beam tracking exceeds the acoustic diffraction limit by an order of magnitude (approximately 2 mm considering the 350 kHz center frequency) and surpasses the physical proton beam spot size typically achieved by current clinical PBT systems (minimum spot size σ = 3.5 mm in air and 7 mm in water for highest-energy beams, measured by Gaussian fitting of the spot profile) [32]. Experiments tracking proton beam trajectories and imaging temporal dose accumulation of M-shaped treatment plans demonstrated that iRABL, operating at a frame rate of 1 kHz (matching the maximum pulse repetition rate of the Proteus® ONE system used in this study), accurately localized individual proton pulses. Using the clinical standard gamma index evaluation with 3mm/3% criteria, the iRABL system met clinical acceptance standards for dose verification.

Super-resolution, high-accuracy, high-sensitivity single-pulse proton spot localization has the potential to transform advanced PBS from a primarily pre-planned technique into a truly image-guided modality by providing pulse-by-pulse, spot-based information on delivered dose location and magnitude with submillimeter precision. PBS is already the leading approach in proton therapy due to its ability to sculpt dose distributions to complex targets with spot scanning, but its clinical accuracy has not been fully validated in humans during PBT due to the lack of clinical tools capable of tracking individual spots, pulses, and delivered doses in real time within patient’s body [33]. The iRABL-based localization approach, offering high single-pulse temporal resolution and high sensitivity to deposited dose without interfering with treatment, provides this missing validation and localization guidance capability. With a sub-diffraction resolution of 0.1 mm, we are providing a level of precision that exceeds the actual physical width of the pencil beam itself. This allows for an incredibly sharp definition of the "Bragg Peak" for proton dose delivery. The 1 kHz frame rate is a “game changer” for PBS, since PBS delivers dose point-by-point and hence a kilohertz capture rate allows verification of every single "spot" in the treatment plan in real time, rather than just the integrated total. A 90% passing rate in gamma analysis at the 3mm/3% criteria is an excellent benchmark for clinical viability, proving that our iRABL system is not just high speed but also highly accurate. The iRABL approach validated in this work is adaptable to other synchrotron- or synchrocyclotron-based PBT systems, as well as to heavy-ion therapy systems such as carbon or helium, which may operate with different pulse repetition rates and durations.

Current clinical concerns about applying PBT more broadly—particularly with advanced techniques such as online adaptation, FLASH radiotherapy, and spatially fractionated therapies—center on range uncertainties from tissue density variations, anatomical changes, interplay effects for moving organs, and the lack of clinically ready tools to localize targets and tissue changes in real time [34-37]. While Monte Carlo simulation and improved stopping power ratio calculations can predict dose distributions with increasing accuracy, validation in human anatomy with clinical beams remains challenging [38]. Conventional *in vivo* dosimetry approaches in PBT face fundamental limitations including image quality of proton radiography, insufficient spatial resolution to resolve submillimeter features, and the need for bulky detector structures or additional shielding to improve signal-to-noise ratios by removing secondary radiation [39-41]. iRABL addresses these limitations by enabling non-invasive, real-time proton spot localization in human body, potentially allowing patient-specific range uncertainties to be assessed during treatment. Furthermore, iRABL’s pulse-by-pulse detection capability could enable dosimetry for emerging techniques that are currently limited by conventional detector technology. FLASH radiotherapy dosimetry is constrained by charge collection and dose-rate effects in ionization chambers and diode detectors. Spatially fractionated proton therapy, such as minibeam-like regimens, requires accurate determination of peak-to-valley dose ratios in narrow beams with steep dose gradients [42-44]. iRABL’s super-resolution *in vivo* dosimetry capability could address both conventional and FLASH beam delivery scenarios, facilitate increasingly precise treatment delivery, and advance PBS toward the vision of "proton surgery."

Despite every exciting and promising results achieved in this study, several limitations should be addressed in future development of iRABL technology. First, the current iRABL system only provides relative dose measurements. To achieve absolute dose measurement capability, a comprehensive calibration protocol is needed that accounts for the signal response of the imaging system, the temporal shape of the radiation pulse, and the tissue properties (e.g., physical density, speed of sound, coefficient of thermal expansion, and specific heat capacity). Specifically, for iRABL, tissue properties vary among individuals but could be measured by the existing imaging methods such as CT, MRI, and ultrasound, and the information could be incorporated into the reconstruction algorithm to further improve the accuracy of beam localization and dose mapping [45-47]. Second, as shown in Figure 6 for the beam trajectory tracking, the proton beam positions tracked by iRABL show higher accuracy especially in the central region of the field of view of the 2D MAT, while the beam tracking at the boundaries of the field of view shows relatively larger discrepancies compared to the treatment plan. Future work will focus on optimizing the design of the 2D MAT to enable a broader field of view, ensuring better consistency when tracking the beam over a large volume within patient’s body. Using a distributed acoustic sensor system instead of a single 2D MAT may offer improved performance and warrant further investigation. Third, the iRABL system provides information regarding the location and the dose of the proton beam without offering anatomical information of patient. To track the relative location of the proton beam within patient’s body and ensure accurate treatment delivery, iRABL should be combined with other anatomical imaging modalities such as MRI, CT, and ultrasound. As demonstrated by several ultrasound based dual-modality imaging setups [48-51], utilizing the same 2D MAT could enable inherently co-registered ultrasound anatomical imaging and iRABL beam localization to achieve beam trajectory tracking and temporal dose mapping on anatomical structures. Being able to map the pencil beam and dose delivery on anatomy, pulse-by-pulse, with submillimeter precision and clinical grade accuracy, the "safety margin" added around the tumor to account for patient’s movement and uncertainty can potentially shrink to near-zero for future iRABL-guided PBT.

In summary, this study developed and validated a super-resolution, high-speed, and high-sensitivity iRABL system capable of localizing the proton beam delivery deep inside a human patient without interrupting treatment. The system enabled proof-of-concept experiments on both prostate cancer patients and tissue-equivalent phantoms, establishing the feasibility of iRABL for clinical application during PBT by tracking spot scanning trajectories and temporal dose accumulation. While performance characteristics such as sensitivity and spatial resolution can be further improved, the iRABL system presented in this work holds great promise for guiding proton beam delivery with submillimeter accuracy, advancing current PBT toward the vision of "proton surgery."

## Acknowledgements

We thank the staff at the University of Florida Health Proton Therapy Institution for cooperating in the patient study. This work was supported by the National Cancer Institute (grant nos. R37CA222215 and R01CA266803).

## Data Availability

All data produced in the present work are contained in the manuscript.

## References

1. Mohan, R., A review of proton therapy–Current status and future directions. Precision radiation oncology, 2022. 6(2): p. 164–176.

2. Miralbell, R., et al., Potential reduction of the incidence of radiation-induced second cancers by using proton beams in the treatment of pediatric tumors. International Journal of Radiation Oncology* Biology* Physics, 2002. 54(3): p. 824–829.

3. Paganetti, H., Range uncertainties in proton therapy and the role of Monte Carlo simulations. Physics in Medicine & Biology, 2012. 57(11): p. R99.

4. Merchant, T.E., et al., Proton versus photon radiotherapy for common pediatric brain tumors: comparison of models of dose characteristics and their relationship to cognitive function. Pediatric blood & cancer, 2008. 51(1): p. 110–117.

5. Pidikiti, R., et al., Commissioning of the world’s first compact pencil-beam scanning proton therapy system. Journal of applied clinical medical physics, 2018. 19(1): p. 94–105.

6. Lane, S.A., J.M. Slater, and G.Y. Yang, Image-guided proton therapy: A comprehensive review. Cancers, 2023. 15(9): p. 2555.

7. Lomax, A., Intensity modulated proton therapy and its sensitivity to treatment uncertainties 2: the potential effects of inter-fraction and inter-field motions. Physics in Medicine & Biology, 2008. 53(4): p. 1043.

8. Pakela, J.M., et al., Management of motion and anatomical variations in charged particle therapy: past, present, and into the future. Frontiers in Oncology, 2022. 12: p. 806153.

9. Li, H., et al., Robust optimization in intensity-modulated proton therapy to account for anatomy changes in lung cancer patients. Radiotherapy and Oncology, 2015. 114(3): p. 367–372.

10. Perkins, G.H., et al., Gliomatosis cerebri: improved outcome with radiotherapy. International Journal of Radiation Oncology* Biology* Physics, 2003. 56(4): p. 1137–1146.

11. Unkelbach, J., et al., Reducing the sensitivity of IMPT treatment plans to setup errors and range uncertainties via probabilistic treatment planning. Medical physics, 2009. 36(1): p. 149–163.

12. Reaz, F., et al., Sharp dose profiles for high precision proton therapy using strongly focused proton beams. Scientific reports, 2022. 12(1): p. 18919.

13. Ku, Y., et al., Tackling range uncertainty in proton therapy: development and evaluation of a new multi-slit prompt-gamma camera (MSPGC) system. Nuclear Engineering and Technology, 2023. 55(9): p. 3140–3149.

14. Knopf, A.-C. and A. Lomax, In vivo proton range verification: a review. Physics in Medicine & Biology, 2013. 58(15): p. R131.

15. Krimmer, J., et al., Prompt-gamma monitoring in hadrontherapy: A review. Nuclear Instruments and Methods in Physics Research Section A: Accelerators, Spectrometers, Detectors and Associated Equipment, 2018. 878: p. 58–73.

16. Paganetti, H. and G. El Fakhri, Monitoring proton therapy with PET. The British journal of radiology, 2015. 88(1051): p. 20150173.

17. Cuneo, K.C., et al., Clinical pilot study of ionizing radiation acoustic imaging (iRAI) for real-time visualization of radiation therapy dose delivery in cancer patients. International Journal of Radiation Oncology* Biology* Physics, 2025.

18. Wang, S., et al., Real-time tracking of the Bragg peak during proton therapy via 3D protoacoustic Imaging in a clinical scenario. npj Imaging, 2024. 2(1): p. 34.

19. Patch, S.K., D. Santiago-Gonzalez, and B. Mustapha, Thermoacoustic range verification in the presence of acoustic heterogeneity and soundspeed errors - Robustness relative to ultrasound image of underlying anatomy. Med Phys, 2019. 46(1): p. 318–327.

20. Parodi, K. and W. Assmann, Ionoacoustics: A new direct method for range verification. Modern Physics Letters A, 2015. 30(17): p. 1540025.

21. Zhang, W., et al., Real-time, volumetric imaging of radiation dose delivery deep into the liver during cancer treatment. Nature biotechnology, 2023. 41(8): p. 1160–1167.

22. Huang, Y., et al., Towards quantitative ionizing radiation acoustic imaging (iRAI) for radiation dose measurement: Validation from simulations to experiments. Medical Physics, 2025. 52(9): p. e18091.

23. Huang, Y., et al., Ionizing radiation acoustic and ultrasound dual-modality imaging for visualization of dose on anatomical structures during radiotherapy. Photoacoustics, 2025: p. 100742.

24. Errico, C., et al., Ultrafast ultrasound localization microscopy for deep super-resolution vascular imaging. Nature, 2015. 527(7579): p. 499–502.

25. Telichko, A.V., et al., Passive cavitation mapping by cavitation source localization from aperture-domain signals—Part I: Theory and validation through simulations. IEEE transactions on ultrasonics, ferroelectrics, and frequency control, 2020. 68(4): p. 1184–1197.

26. Telichko, A.V., et al., Passive cavitation mapping by cavitation source localization from aperture-domain signals—Part II: Phantom and in vivo experiments. IEEE transactions on ultrasonics, ferroelectrics, and frequency control, 2020. 68(4): p. 1198–1212.

27. Zhao, L., et al., Developing an accurate model of spot-scanning treatment delivery time and sequence for a compact superconducting synchrocyclotron proton therapy system. Radiation Oncology, 2022. 17(1): p. 87.

28. Oku, Y., et al., Analysis of suitable prescribed isodose line fitting to planning target volume in stereotactic body radiotherapy using dynamic conformal multiple arc therapy. Practical Radiation Oncology, 2012. 2(1): p. 46–53.

29. Park, J.M., et al., Reliability of the gamma index analysis as a verification method of volumetric modulated arc therapy plans. Radiation oncology, 2018. 13(1): p. 175.

30. Zhang, W., et al., Real-time, volumetric imaging of radiation dose delivery deep into the liver during cancer treatment. Nature Biotechnology, 2023: p. 1–8.

31. Zhang, W., et al., Dual-Modality X-Ray-Induced Radiation Acoustic and Ultrasound Imaging for Real-Time Monitoring of Radiotherapy. BME Frontiers, 2020. 2020: p. 9853609.

32. Ricci, J.C., et al., The root cause analysis on failed patient-specific measurements of pencil beam scanning protons using a 2D detection array with finite size ionization chambers. Journal of Applied Clinical Medical Physics, 2021. 22(8): p. 175–190.

33. Parodi, K., Latest developments in in-vivo imaging for proton therapy. The British journal of radiology, 2020. 93(1107): p. 20190787.

34. Sterpin, E., et al., Robustness evaluation of pencil beam scanning proton therapy treatment planning: A systematic review. Radiotherapy and Oncology, 2024. 197: p. 110365.

35. Kang, M., et al., A study of the beam-specific interplay effect in proton pencil beam scanning delivery in lung cancer. Acta Oncologica, 2017. 56(4): p. 531–540.

36. Nesteruk, K.P. and S. Psoroulas, FLASH irradiation with proton beams: beam characteristics and their implications for beam diagnostics. applied sciences, 2021. 11(5): p. 2170.

37. Bookbinder, A., et al., Validation and reproducibility of in vivo dosimetry for pencil beam scanned FLASH proton treatment in mice. Radiotherapy and Oncology, 2024. 198: p. 110404.

38. Niepel, K., et al., Validation of dual-energy CT-based composition analysis using fresh animal tissues and composition-optimized tissue equivalent samples. Physics in Medicine & Biology, 2024. 69(16): p. 165033.

39. Johnson, R.P., Meeting the detector challenges for pre-clinical proton and ion computed tomography. Physics in Medicine & Biology, 2024. 69(11): p. 11TR02.

40. Moglioni, M., et al., In-vivo range verification analysis with in-beam PET data for patients treated with proton therapy at CNAO. Frontiers in Oncology, 2022. 12: p. 929949.

41. Pinto, M., Prompt-gamma imaging in particle therapy. The European Physical Journal Plus, 2024. 139(10): p. 884.

42. Schneider, T., et al., Advancing proton minibeam radiation therapy: magnetically focussed proton minibeams at a clinical centre. Scientific reports, 2020. 10(1): p. 1384.

43. Mohan, R. and D. Grosshans, Proton therapy–present and future. Advanced drug delivery reviews, 2017. 109: p. 26–44.

44. Yang, F., et al., Proton minibeam radiotherapy: a review. Frontiers in Oncology, 2025. 15: p. 1580513.

45. Webb, T.D., et al., Measurements of the relationship between CT Hounsfield units and acoustic velocity and how it changes with photon energy and reconstruction method. IEEE transactions on ultrasonics, ferroelectrics, and frequency control, 2018. 65(7): p. 1111–1124.

46. Johnson, S.L., et al., Development and validation of a MRgHIFU non-invasive tissue acoustic property estimation technique. International Journal of Hyperthermia, 2016. 32(7): p. 723–734.

47. Glozman, T. and H. Azhari, A method for characterization of tissue elastic properties combining ultrasonic computed tomography with elastography. Journal of Ultrasound in Medicine, 2010. 29(3): p. 387–398.

48. Huang, Y., et al., Ionizing radiation acoustic and ultrasound dual-modality imaging for visualization of dose on anatomical structures during radiotherapy. Photoacoustics, 2025. 44: p. 100742.

49. Oraiqat, I., et al., An ionizing radiation acoustic imaging (iRAI) technique for real-time dosimetric measurements for FLASH radiotherapy. Medical physics, 2020. 47(10): p. 5090–5101.

50. Kim, J., et al., Enhanced dual-mode imaging: Superior photoacoustic and ultrasound endoscopy in live pigs using a transparent ultrasound transducer. Science advances, 2024. 10(47): p. eadq9960.

51. Yu, Y., et al., Simultaneous photoacoustic and ultrasound imaging: A review. Ultrasonics, 2024. 139: p. 107277.

